# Cerebrospinal fluid total tau levels indicate aberrant neuronal plasticity in Alzheimer’s disease

**DOI:** 10.1101/2020.10.29.20211920

**Authors:** Pieter Jelle Visser, Lianne M. Reus, Johan Gobom, Iris Jansen, Ellen Dicks, Magda Tsolaki, Frans R.J. Verhey, Julius Popp, Pablo Martinez-Lage, Rik Vandenberghe, Alberto Lleó, José Luís Molinuevo, Sebastiaan Engelborghs, Yvonne Freund-Levi, Lutz Froelich, Kristel Sleegers, Valerija Dobricic, Shengjun Hong, Simon Lovestone, Johannes Streffer, Stephanie J.B. Vos, Isabelle Bos, ADNI, August B. Smit, Kaj Blennow, Philip Scheltens, Charlotte E. Teunissen, Lars Bertram, Henrik Zetterberg, Betty M. Tijms

## Abstract

Alzheimer’s disease (AD) is characterised by abnormal amyloid beta and tau processing. Previous studies reported that cerebrospinal fluid (CSF) total tau (t-tau) levels vary between patients. Here we show that CSF t-tau variability is associated with distinct impairments in neuronal plasticity mediated by gene repression factors SUZ12 and REST. AD individuals with abnormal t-tau levels have increased CSF concentrations of plasticity proteins regulated by SUZ12 and REST. AD individuals with normal t-tau, on the contrary, have decreased concentrations of these plasticity proteins and increased concentrations in proteins associated with blood-brain and blood CSF-barrier dysfunction. Genomic analyses suggested that t-tau levels in part depend on genes involved in gene expression. The distinct plasticity abnormalities in AD as signaled by t-tau urge the need for personalised treatment.

---

According to the amyloid cascade hypothesis, AD starts with amyloid beta (Aβ) aggregation followed by tau pathology (*1*). Between onset of Aβ aggregation and dementia, the disease has a preclinical stage of 10 years in which cognition is normal and a prodromal stage of 4 years with mild cognitive impairment (*2*). Increased CSF total tau (t-tau) levels are supposed to reflect axonal loss and disease severity, as tau stabilizes axonal microtubules (*3*). Still, CSF t-tau is abnormal in 50% of preclinical AD individuals, when neurodegeneration is limited (*4*), and normal in 25% of individuals with prodromal and dementia AD (*5,6*). An alternative explanation of variability in CSF t-tau levels may be differences in synaptic activity and neuronal plasticity (*7,8*). As tau-related mechanisms are emerging as new treatment targets for AD, it is crucial to understand drivers of CSF tau dysregulation. We hypothesized that differences in CSF t-tau levels are associated with neuronal plasticity markers, in particular in early AD stages.

We performed a proteomic, genomic and imaging study including 1380 individuals in the AD continuum (*9*), defined by abnormal CSF Aβ_1-42_, in the preclinical (n=280), prodromal (n=729) and dementia stage (n=371) and 380 controls with normal cognition and normal CSF Aβ_1-42_ and t-tau from the Alzheimer’s Disease Neuroimaging Initiative (ADNI) and the EMIF-AD Multimodality Biomarker Discovery study (*3*). Of the individuals with AD, 788 (57%) had abnormally high CSF t-tau levels and 592 normal t-tau levels. Abnormal and normal t-tau groups showed generally similar demographics and baseline cognitive performance (sTable 1a).

We first determined stability of t-tau levels over time in a subset of 499 ADNI participants with repeated CSF sampling up to 7 years. CSF t-tau increased in each clinical stage but this increase did not differ between abnormal and normal t-tau groups, and also not from controls (sTable2b, sFigure 1a). Tau status at last measurement (on average 2.4 years after baseline), remained normal in 80% individuals with initially normal t-tau and those who became abnormal scored at baseline just below the cut-point (sTable 2b, sFigure 1b). This suggests that CSF t-tau levels reflect a trait rather than a stage.

**Figure 1.**
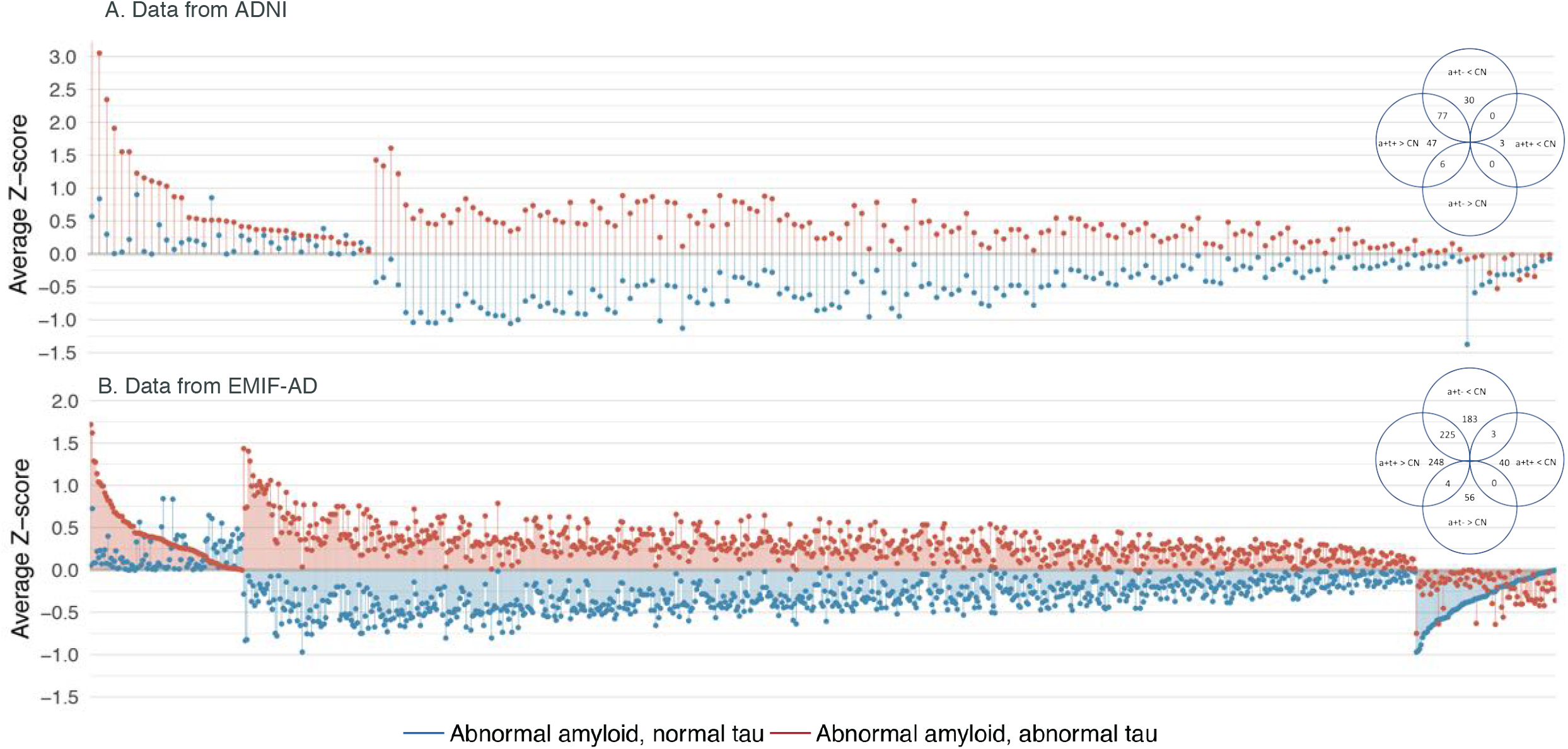
Protein concentrations relative to control group in individuals with AD according to t-tau status. Protein concentrations of individuals with abnormal Aβ_1-42_ and abnormal total tau (a+t+, red) and abnormal Aβ_1-42_ and normal t-tau (a+t-, blue) in ADNI (A) and EMIF-AD (B). Concentrations are expressed as z-score relative to control group with normal cognition, Aβ_1-42_ and t-tau (z-score=0). Proteins are sorted according to change relative to control group. Shown are proteins that differed from controls in either a+t+ or a+t-. Venn diagram shows number of proteins that differed. Full data are shown in sTable2c.

We then studied whether the CSF proteome differed for t-tau subgroups in 559 individuals with proteomic data (sTable 1b). This included targeted proteomics of 248 proteins in ADNI and targeted and untargeted proteomic measures of 1458 proteins in EMIF-AD (sTable 2c). Compared to controls, AD individuals with abnormal CSF t-tau had increased levels of 33% (ADNI) to 52% (EMIF-AD) of the proteins measured, and decreased levels of 1 to 3% proteins (figure 1, sTable 2a,c). AD individuals with normal CSF t-tau, showed an opposite pattern with increased levels of 3 to 4% of the proteins and decreases in 28% (ADNI) to 43% (EMIF-AD) of the proteins. Around 50% of the proteins increased in individuals with abnormal t-tau were decreased in individuals with normal t-tau in both cohorts. This indicates that both t-tau groups show disruptions in the same molecular processes, but in opposite directions. Enrichment analyses using the Gene Ontology biological processes (GO-BP) for CSF proteins with increased levels in AD individuals with abnormal t-tau relative to controls, showed involvement of neuronal plasticity-related processes such as nervous system development, axongenesis, synapse assembly, myelination, gliogenesis, angiogenesis, MAPK signaling, cell-cycle, gene expression and glycolysis (Figure 2; sTable 3a shows all biological processes; Table 3b and sFigure 2 show synapse related processes). Abnormality in each of these processes has previously been reported in AD (*10-15*), and our proteomic findings now show that they are all part of an aberrant plasticity response. Increased proteins further showed involvement of cytokine signaling, leukocyte activation, oxidative stress, mitochondrial dysfunction and apoptosis. Amyloid production was increased as well, reflected by increased levels of amyloid precursor protein (APP), Aβ_1-40_, Aβ_1-38,_ increased levels and activity of beta-site amyloid precursor protein cleaving enzyme 1 (BACE1), and increased concentrations in substrates of BACE1, alpha secretase and gamma secretase, the main secretases involved in Aβ metabolism (sTable2c). Increased Aβ_1-42_ production is known to set off the amyloid cascade in autosomal dominant AD (*16*), and our data suggest that increased amyloid production may also play a role in sporadic AD. Increased Aβ_1-42_ production is related to neuronal plasticity because APP and oligomeric Aβ_1-42_ activate mitogenic MAPK/ERK signaling, and the APP intracellular domain (AICD) stimulates gene transcription through interaction with APBB1(*13,17,18*). To identify other potential drivers of increased protein levels we performed CHeA transcription factor-binding site enrichment analysis, which indicated SUZ12 (120 proteins, p-FDR corrected=1.62E-11) and REST (92 proteins, p-FDR corrected =1.04E-9) as most significantly enriched. SUZ12 and REST repress gene transcription through H3 histone acetylation, and play a role in neuronal development and neurodegeneration (*19,20*). SUZ12 and REST were previously associated with increased expression of plasticity genes in iPSC neuronal progenitor cells of sporadic AD individuals, of which 63 proteins were also increased in CSF in our study in individuals with abnormal Aβ_1-42_ and t-tau (*15*). REST has also been associated with the increased expression of plasticity related genes in tangle bearing neurons (*21*). Tau may increase gene expression directly or indirectly by increasing H3K9 acetylation, REST translocation or chromatin relaxation (*15,22,23*). In summary, the CSF proteome of AD individuals with abnormal t-tau reflects a disbalance of mitogenic signaling and repressive transcriptional activity leading to increased gene expression of a wide range of neuronal plasticity proteins. As this may also further increase expression of APP and t-tau, this could induce a vicious cycle.

**Figure 2.**
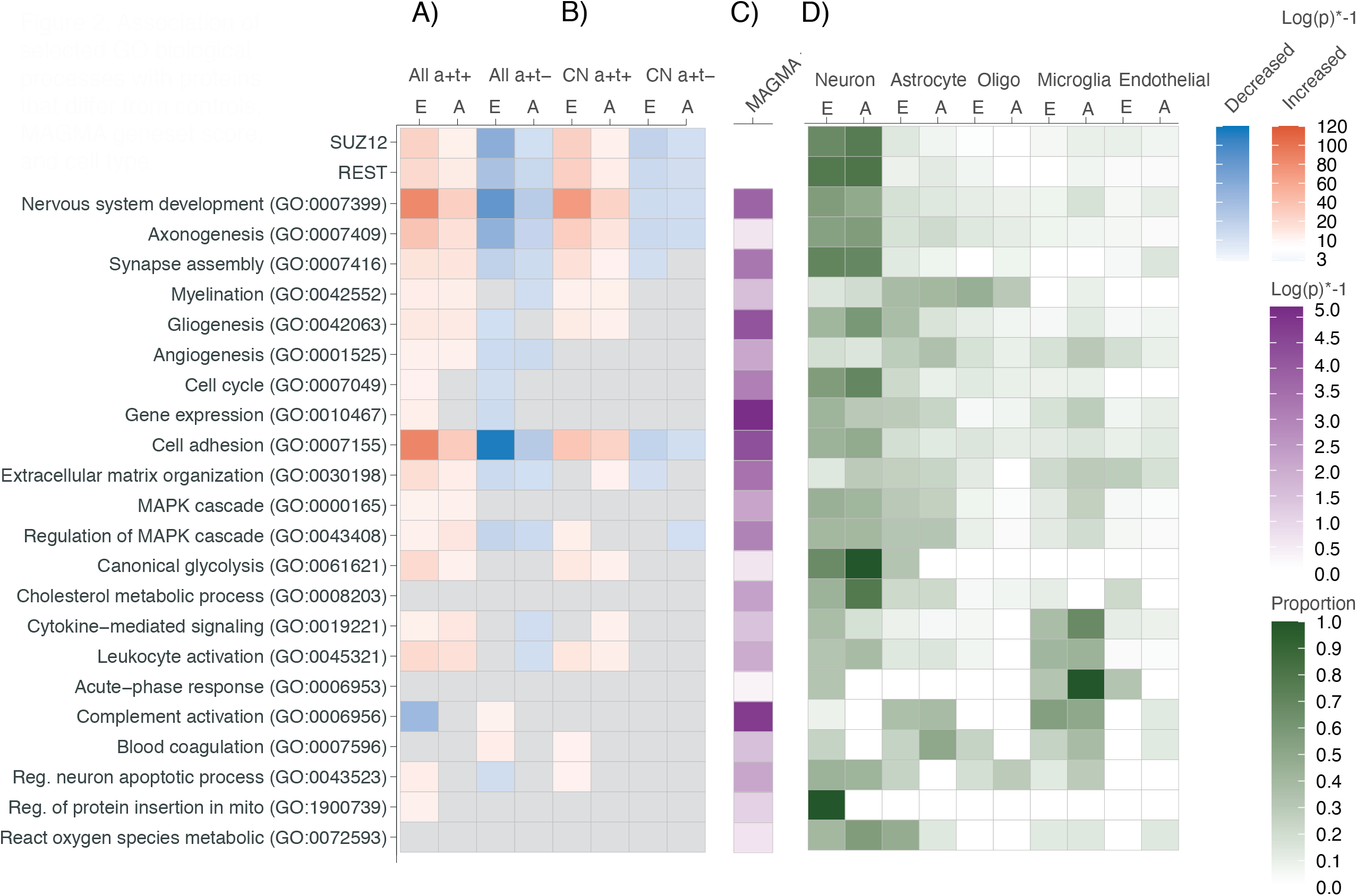
Association of selected GO biological processes with proteins that differ from controls, MAGMA geneset score, and cell type. Heatmap showing p-value (log p-value*-1) of selected enriched GO biological processes and SUZ12 and REST transcription factors of proteins increased (red) or decreased (blue) relative to controls in total sample (A) or in preclinical stage (B); P-value of association of GO biological processes with abnormal t-tau in GWAS-based MAGMA genset analysis (C); Proportion of proteins with cell-type specific expression (D). Data are shown for ADNI (A) and EMIF-AD (E) separately as indicated. Data used in heatmap are shown in sTable 7. P-values of all GO biological processes are listed in sTable 3a (CSF enrichment) and sTable 4D (MAGMA analysis). A+t+: abnormal Aβ_1-42_ and t-tau; A+t-: abnormal Aβ_1-42_ and normal total tau; CN A+t+: abnormal Aβ_1-42_ and t-tau in preclinical stage; CN A+t-: abnormal Aβ_1-42_ and normal t-tau in preclinical stage. Oligo= oligodendrocyte; endothelial=endothelial cell.

In AD individuals with normal t-tau, the processes related to neuronal plasticity in particular were abnormally decreased with concomitant lower levels of APP, Aβ_1-40_ and Aβ_1-38_, and secretase substrates (Figure 2). Proteins with decreased concentrations also converged on SUZ12 and REST transcription factors, suggesting that increased gene repression activity is driving this CSF proteomic profile. Increased repressive activity of REST and SUZ12 has been reported in brain ischemia, seizures, and toxic insults supposedly reducing excitotoxicity (*19,20,24-26*). Among the 67 proteins with increased levels relative to controls, 20 proteins were associated with blood-brain barrier (BBB) dysfunction (*27*). They also included 25 proteins with high expression in choroid plexus (CP), and this CP overreprensentation was confirmed by ABAEnrichment analysis (minimum pFWER=0.004), which could suggest blood-CSF barrier (BCSFB) dysfunction. Increased proteins were mainly seen in the EMIF-AD cohort, as ADNI proteomics was targeted for brain specific proteins. Impaired BBB/BCSFB function may result from toxic effects of Aβ on cells that constitute these barriers (*28*). Barrier dysfunction reduces neuronal plasticity through impaired glucose transport, capillary perfusion, and neurogenesis (*28*), which is possibly reflected in the hypoplasticity signal we observed in CSF. When we repeated analysis by comparing CSF protein concentrations between AD individuals with normal and abnormal t-tau we found enrichment for the same processes that differed for each group relative to controls (sTable2c, sFigure 3a). Analysis of cell-type specific proteins showed that plasticity proteins were strongly associated with neuron-specific proteins (Figure 2, sTable 2d). Stratified analyses for disease stage further showed that alternations in CSF proteome related to neuronal plasticity were already present in the preclinical stage (Figure 2, sTable 2c, e), and in both groups decreased with disease stage (sTable 2c, Figure 3, sFigure 3b). In the preclinical stage of individuals with normal t-tau levels, proteins associated with choroid plexus were increased (sTable 2e) and with increasing disease severity concentrations of BBB related proteins increased (Figure 3).

**Figure 3.**
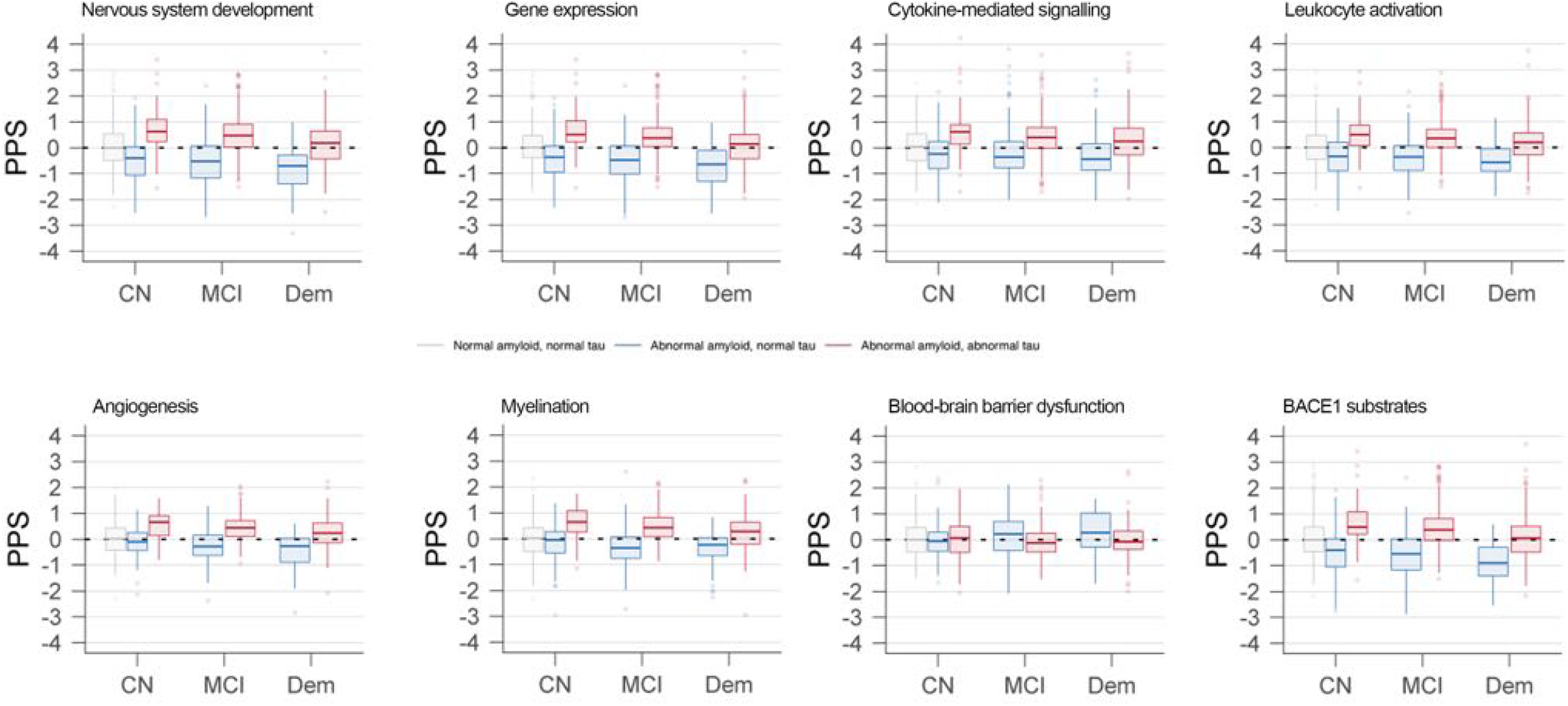
Association of protein process score (PPS) with disease severity. Data are based on cross-sectional analysis and combines data from ADNI and EMIF-AD. In grey PPS of control group with normal cognition and normal CSF Aβ_1-42_ and t-tau; In red PPS of individuals with abnormal Aβ_1-42_ and t-tau; In blue PPS of individuals with abnormal Aβ_1-42_ and normal t-tau. CN=normal cognition; MCI=mild cognitive impairment; Dem=dementia.

We next aimed to identify genetic factors related to increased t-tau in AD individuals in the combined ADNI and EMIF-AD cohorts (n=1067). *APOE* ε4, the major genetic AD risk factor, was more common in the abnormal t-tau group (66% vs 53% p<0.001), and in both AD groups *APOE* ε4carriership was more common than in controls (17%, p<0.001, sTable 1a). Compared to controls, both abnormal and normal t-tau groups showed higher AD polygenic risk scores (PGRS) across SNP inclusion thresholds (Figure 4, sTable4a) (*29*). In none of the clinical stages, PGRS differed between AD individuals with or without increased t-tau after correction for *APOE* ε4 carriership and age (sTable 4a). Thus, both groups have a similar presence of AD genetic risk variants. An exploratory GWAS within the AD group to identify potential genetic markers associated with abnormal t-tau in EMIF-AD and ADNI (combined by meta-analysis), showed tentative associations with SNPs in the *APBB2* gene (p-values 1.63-2.92E10-6, sTable 4b), which previously were associated with AD (*30*). *APBB2* binds AICD and its overexpression has been associated with increased levels of Aβ_1-40_ and AICD (*31*). The top 3 genes associated with abnormal t-tau from MAGMA gene-based analyses showed in addition to *ABPP2, TBC1D10B* (p=2.9E-04), a Rab GTPA activating protein involved in MAPK signaling, and *LRP3* (p=3.36E-04), a low-density lipoprotein receptor protein (sTable 4c). The top 3 GO-BP gene-sets associated with abnormal t-tau from MAGMA were positive regulation of cellular process (self-contained p=1.07E-04), protein K29-linked ubiquitination (p= 1.52E-04) and negative regulation of actin nucleation (p=1.57E-04, sTable 4d). Twenty-eight percent of GO-BPs associated with proteins that differed between AD individuals with abnormal and normal t-tau also differed between tau groups in MAGMA gene-set analysis (sTable 3a), suggesting that differences in CSF t-tau concentration have partly a genetic basis. Then we directly correlated CSF protein concentrations with subject level genetic risk scores (GRS) for the top 3 genes and GO-BP and 9 GO-BPs listed in figure 2 that differed between abnormal and normal t-tau groups in both CSF and MAGMA geneset analysis. Given the association of CSF profiles with REST and SUZ12 we also selected 2 histone acetylation GO-BPs with MAGMA self-contained p-value<0.05. The strongest positive correlations between genetic risk scores with CSF proteins were observed for gene expression (288 proteins), regulation of MAPK cascade (146 proteins), and histone H3 acetylation (127 proteins, sTable 5, figure 5). This further supports a genetic basis of t-tau differences between groups.

**Figure 4.**
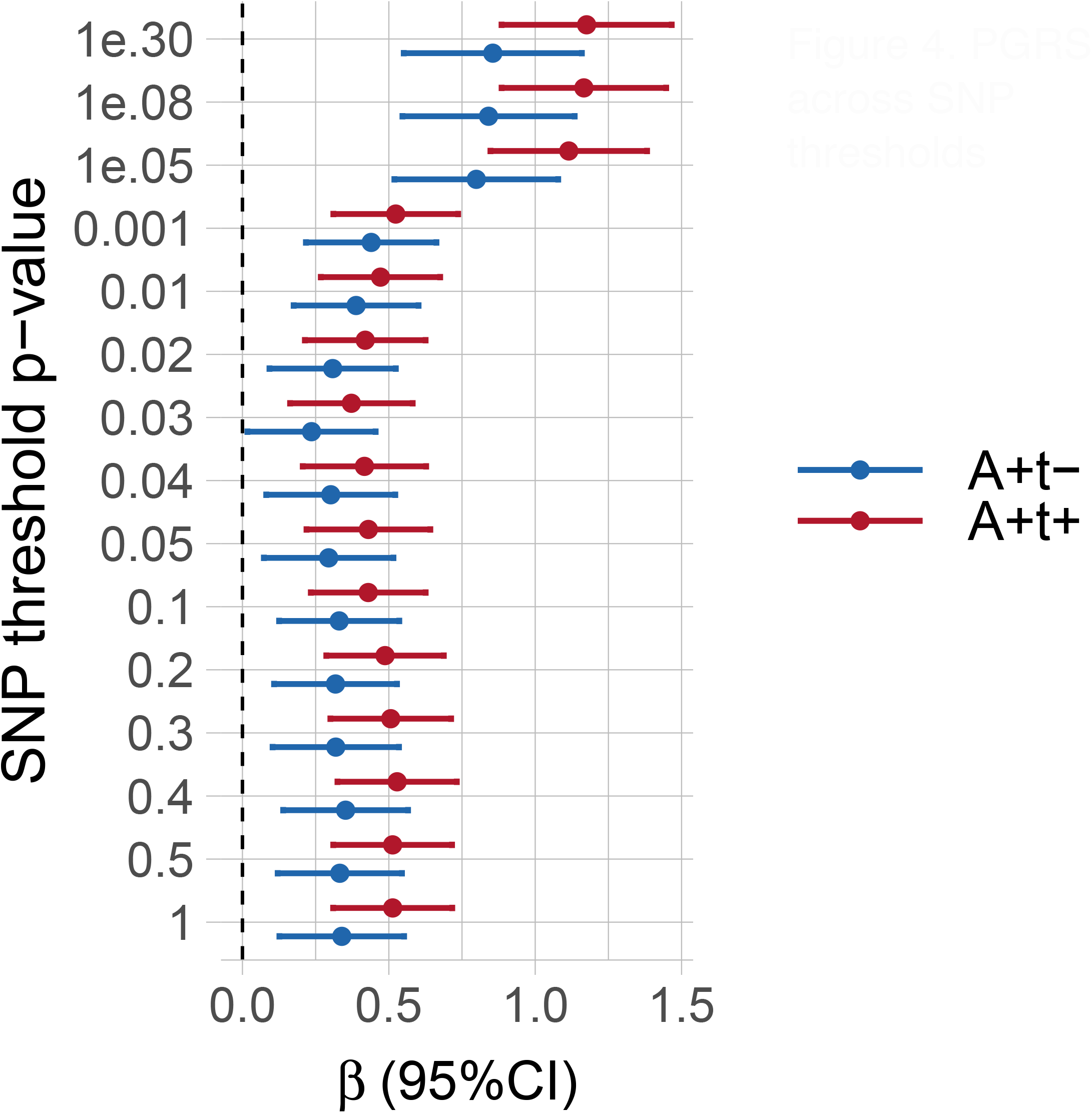
PGRS across SNP thresholds. SNP weights were based on AD GWAS (Jansen et al 2019). Full data (including data after correction for APOE ε4 genotype and demographics, and separate for cohort and clinical stage) are shown in sTable 4a. A+t+: abnormal Aβ_1-42_ and t-tau (red); A+t-: abnormal Aβ_1-42_ and normal t-tau (blue).

**Figure 5.**
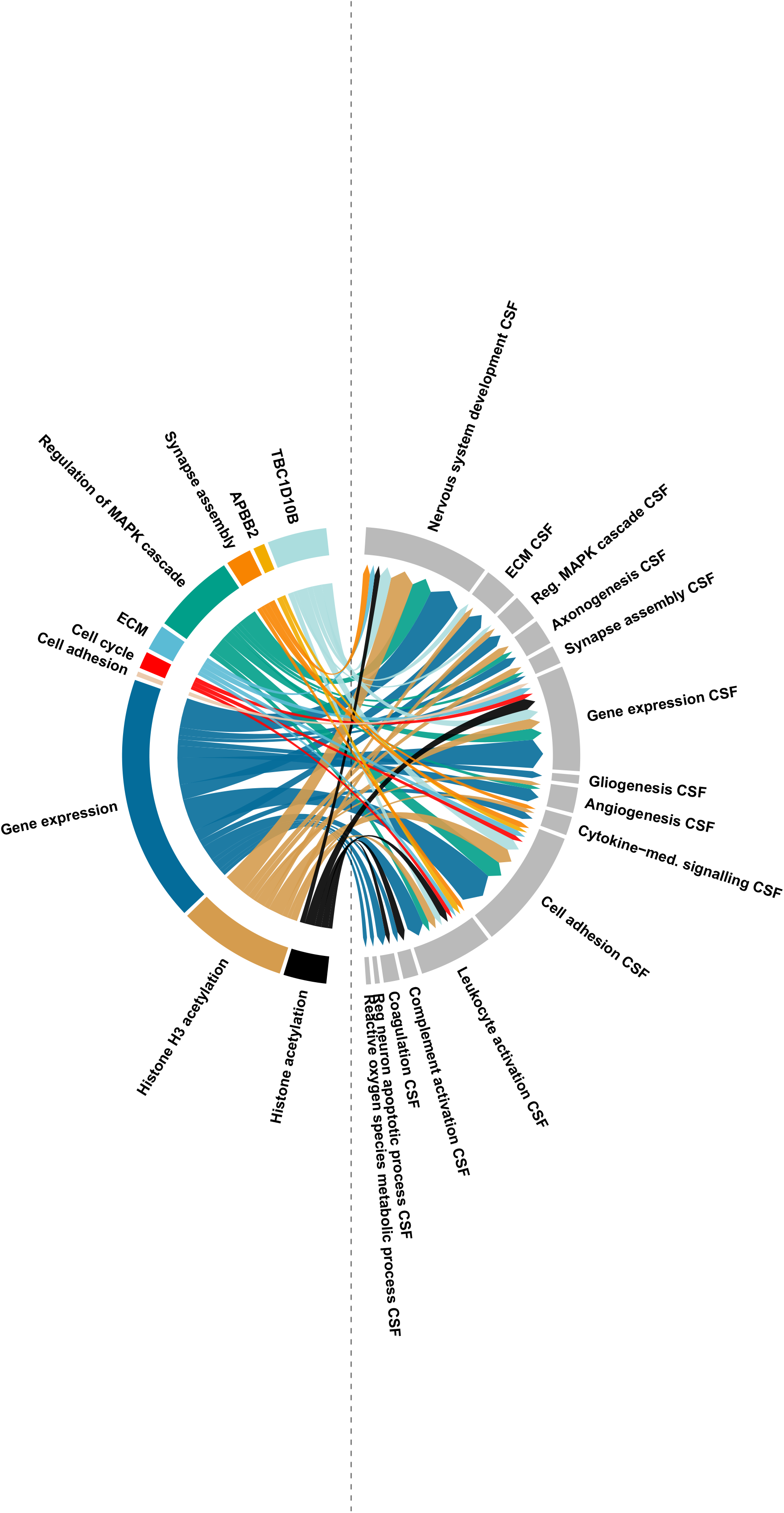
Relation between genetic risk score and CSF protein correlation. Relation between genetic risk score and CSF protein concentration. Shown are the 10 GRS from genes or GO-biological process (GO-BP) with the highest number of positive correlations with CSF proteins. The size of the arrows visually indicates how many proteins that correlate with the GRS in the coloured left outer ring belong to one of the CSF GO-BP shown in the right grey outer ring. For example, the GRS of GO-BP gene expression showed a positive correlation with 256 CSF proteins and these proteins were predominantly associated with GO-BP of nervous system development, gene expression and cell adhesion. Only arrows to CSF GO-BP are shown in case the GRS correlated with at least 8 proteins from the CSF GO-BP. Supporting data in sTable 5. ECM= Extracellular matrix.

Finally, we studied effects of t-tau on disease progression using longitudinal ADNI data. Relative to controls, both preclinical AD groups showed faster cognitive decline and PET-amyloid accumulation (Figure 6, sFigure 4 and 5, sTable 6). Compared to individuals with AD and normal t-tau, individuals with abnormal t-tau declined faster on cognitive tests, cortical thickness and glucose metabolism in prodromal AD and showed faster amyloid aggregation on PET in all stages. This faster accumulation of Aβ may result from hyperplasticity-related Aβ production.

**Figure 6.**
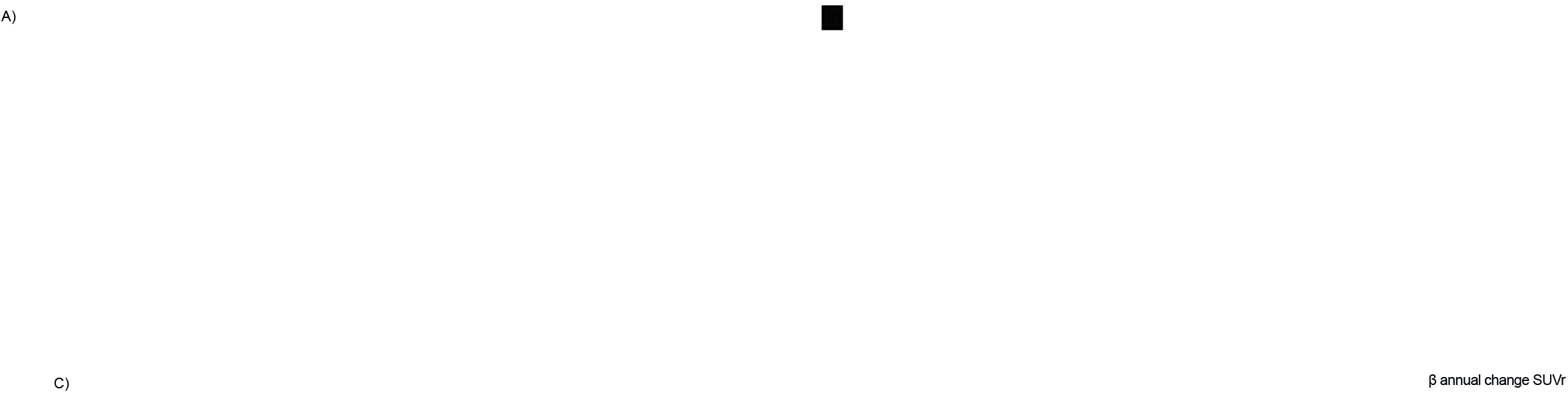
Longitudinal change in cognition and amyloid PET binding. Longitudinal change in Logical memory (A), CDR sum of boxes (B) and PET amyloid aggregation (C) according to diagnostic group and clinical stage. PET amyloid data are not shown for dementia with abnormal Aβ1-42 and normal t-tau because of low number of participants (n=6). In figure 6A and 6B change of controls are shown in grey, of individuals with a+t+ in red, and of individuals with a+t-in blue. Data are from ADNI only. Data are shown in sTable 6a (Logical memory, CDR-sum of boxes) and 6b (amyloid aggregation). CN=normal cognition; MCI=mild cognitive impairment; a+t+= abnormal Aβ_1-42_ and t-tau; a+t-= abnormal Aβ_1-42_ and normal t-tau.

In summary, CSF t-tau levels in AD are related to aberrant neuronal plasticity and dysfunction of the blood-brain and blood-CSF barriers. CSF proteomic analysis indicated that both AD individuals with abnormal and normal t-tau have disturbed mitogenic MAPK signaling and SUZ12/REST signaling, but in opposite ways. Furthermore, opposite CSF proteomic patterns were already observed in the preclinical stage, and persisted in prodromal and mild dementia stages, indicating that they are traits rather than a reflection of disease state.

Our exploratory GWAS, MAGMA analyses and the association between gene risk scores and CSF protein correlations suggest that t-tau levels in part depend on genes involved in gene expression. Larger samples are needed to confirm these results. This genetic pattern is different from that of AD risk genes, which are associated with lipid metabolism and immune activation (*29*). Possibly AD risk genes drive amyloid aggregation, and other genes determine whether an individual will follow a hypo- or hyperplastic trajectory.

Our finding that increased CSF t-tau was associated with increased levels of neuronal plasticity proteins provides further support for the role of tau in neuronal plasticity (*21,23*). Tau may influence gene expression itself (*21-23*), which may explain why knock-out of tau in AD mice models prevented hyperactivation (*32,33*).

A clinical implication of our results is that in the diagnostic framework for AD t-tau should be considered also a marker of dysregulated plasticity (*9*). Treatments for hyperexcitation and hyperplasticity, such as histone modification and anti-epileptic drugs, which are currently tested in clinical trials, may be only beneficial for AD individuals with abnormal t-tau before dementia onset only. AD individuals with normal t-tau and early barrier dysfunction would benefit the most from treatments focusing on prevention of barrier damage and may response differently on antibody therapy than AD individuals with increased t-tau.

## 2. Methods

### 2.1 Study participants

We selected individuals with AD pathology in CSF and a control group with normal cognition and normal AD biomarkers from the Alzheimer’s Disease Neuroimaging Initiative (ADNI, n=961, <u>adni.loni.usc.edu</u>) and the EMIF-AD multimodality biomarker discovery (MBD) study (n=799) (*34*). ADNI started in 2003 as a public-private collaboration under the supervision of Principle Investigator Michael W. Weiner, MD. The primary goal of ADNI is to study whether serial magnetic resonance imaging (MRI), positron emission tomography (PET), other biological markers, and clinical and neuropsychological measures can be combined to measure the progression of mild cognitive impairment (MCI) and early Alzheimer’s disease (AD) and has enrolled 2850 individuals so far, see www.adni-info.org for the latest information. The EMIF-AD MBD study aimed to identify novel markers for diagnosis and prognosis of predementia AD. It combined existing clinical data, samples and scans of 1218 individuals with normal cognition, MCI or mild dementia from prospective cohort studies (*34*). The study collected baseline clinical and neuropsychological data, MRI scans, and plasma, DNA and CSF samples and follow-up diagnosis and cognitive scores (*34*). The institutional review boards of all participating institutions approved the procedures for this study. Written informed consent was obtained from all participants or surrogates.

### 2.2 Group definition and staging

We selected individuals with AD pathology defined as abnormal CSF abeta 1-42 (Aβ_1-42_), and we subdivided this group into abnormal and normal CSF total tau (t-tau) groups (see below for CSF analysis). Based on their cognitive performance, AD individuals were classified in 3 clinical stages as preclinical AD (normal cognition), prodromal AD (mild cognitive impairment, i.e., MCI) and mild to moderate AD-type dementia according to study specific criteria (*34,35*). The control group consisted of individuals with normal cognition, as well as normal CSF Aβ_1-42_ and t-tau levels.

### 2.3 Clinical assessment

As a measure of global cognition we used the Mini-Mental State Examination (MMSE) (*36*) and the ADAS-Cog 11 item version (*37*) (ADNI only). As a measure of memory function we used the delayed recall of the logical memory subscale II of the Wechsler Memory Scale delayed recall (ADNI) (*38*) or a center specific verbal word learning task (EMIF-AD) (*34*). We selected the Clinical Dementia Rating (CDR) scale sum of boxes score as a measure of disease severity (*39*).

### 2.4 CSF analysis

CSF samples were obtained as previously described (*34,40*).

#### Abeta42 and tau measurements

In EMIF-AD CSF Aβ_1-42_ and t-tau were measured locally with INNOTEST ELISA or INNOBIA AlzBio3 INNOBIA AlzBio3 (Fujirebio, Ghent, Belgium) and in ADNI on the xMAP Luminex platform (Luminex Corp, Austin, TX) at the ADNI Biomarker Core laboratory at the University of Pennsylvania Medical Center. Abnormal Aβ_1-42_ and t-tau were defined according to published cut-offs in ADNI (*41*). Cut-offs for Aβ_1-42_ and t-tau were cohort-specific in EMIF-AD (*34*). Because EMIF-AD centers had used different approaches to determine Aβ_1-42_ cut-offs, which could lead to bias, we determined for each cohort specific cut-offs using unbiased Gaussian mixture modelling (sTable 1c) (*42*). We used longitudinal t-tau levels from ADNI (*40*), measured within the same batch within individuals, to assess changes over time.

#### Proteomic analysis ADNI

Details on protein measurements can be found at http://adni.loni.usc.edu/data-samples/biospecimen-data/. YKL-40, neurogranin, soluble APP beta, neurogranin, neurofilament light, alpha synuclein, phosphorylated alpha synuclein, HBB, CFH, sTREM2, VILIP-1, and BACE1 activity were measured by ELISA or related assays. 190 analytes were measured using the multiplex Human DiscoveryMAP panel from Rules Based Medicine (MAP-RBM) (*43*). This panel assesses proteins related to cytokines, chemokines, metabolic markers, hormones, growth factors, tissue remodeling proteins, angiogenesis markers, acute phase reactants, cancer markers, kidney damage markers, and CNS biomarkers. In addition, mass-spectroscopy measures of isoprostanes, Aβ_1-38_, and Aβ_1-40_ (*44*), and 567 peptides representing 222 proteins were performed (the “Biomarkers Consortium CSF Proteomics MRM data set”) (*43,45*). Proteins and peptides were selected based upon their previous detection in CSF, relevance to AD, and results from the MAP-RBM. We used the quality checked and finalised ‘Normalized Intensity’ data as described in the “Data Primer” document at https://adni.loni.usc.edu/wp-content/uploads/2012/01/2011Dec28-Biomarkers-Consortium-Data-Primer-FINAL1.pdf

#### Proteomic analysis EMIF-AD

Neurogranin, neurofilament-light, abd YKL-40, abeta38 and abeta40 were measured by ELISA and Aβ_38_ and Aβ_40_, using the V-PLEX Plus Ab Peptide Panel 1 (6E10) Kit from Meso Scale Discovery (MSD, Rockville, MD) (*3*). Mass spectrometry was performed using tandem mass tag (TMT) technique with 10+1 plexing as previously described, using high-pH reverse phase HPLC for peptide prefractionation (*46*) (Tijms et al in press).The EMIF-AD mass

spectrometry proteomics data have been deposited to the ProteomeXchange Consortium via the PRIDE partner repository with the dataset identifier 10.6019/PXD019910 (*47*). Normalised abundances with associated clinical data can be requested from the EMIF-AD consortium (*3*). Peptides were mapped to 2535 proteins using the software ProteomeDiscoverer v.2.2. (Thermo Scientific), using Mascot (MatrixScience) for protein identification (precursor Dm tolerance 15 ppm, fragment tolerance 0.05 Da, max missed cleavage sites 2), searching the human subset of the UniProtKB SwissProt database (www.uniprot.org). Percolator (MatrixScience) was used for scoring peptide specific matches, and a strict 1% false discovery rate (FDR) was set as threshold for identification. For reporter ion quantification the following settings were used: Integration tolerance 20 ppm; Integration Method Most Confident Centroid; normalize on the reference protein average. The median (IQR) CV for these proteins was 5.6 (3.8, 8.0).

#### Protein selection

Proteins and protein fragments (ADNI) values were normalised according the mean and standard deviation values of the control group. For ADNI, we averaged peptides that mapped to the same protein into a composite protein score when they correlated with r >.5, and included as a single peptide otherwise. When the same protein was measured by different platforms in ADNI, values were averaged if they correlated with r>.5 and otherwise we included them as separate proteins. From the EMIF-AD MRM analysis proteins were included when observed in at least 92 individuals (30% of EMIF-AD sample). For related proteins that had identical values (e.g. ACTA1 and ACTA2), due to fragment a-specificity we randomly selected one protein for analysis.

### 2.5 Proteomic annotation

#### Biological process

We performed pathway enrichment analysis for Gene Ontology biological processes for proteins that differed between groups using the online Panther application (*48*) using FDR corrected Fisher exact tests.

#### Predominant cell-type of protein production

Based on the BRAIN RNASeq database (http://www.brainrnaseq.org) (*49*), we labelled proteins as being predominantly produced by a certain cell type when levels were higher than 40% of old individuals produced across cell types.

#### Transcription factor enrichment

We performed transcription factor enrichment analysis using ChIP-X ChEA through Enrichr and report on adjusted p-values (*50*). <u>SynGO:</u> We mapped proteins that differed between a+t+ and a+t-to location at the synapse using SynGO 1.0 (*51*).

#### Other annotations

- Proteins associated with choroid plexus expression: High expression of proteins in the choroid were based on and the Allen Brain Atlas (*52*) through Harmonizome (*53*) and ABAEnrichment analysis (*54*);
- Blood-Brain barrier associated proteins: Proteins mentioned in (*27,55,56*);
- BACE1 substrates: Proteins mentioned in one of the following articles (*57,58*);
- Alpha secretase substrates: Proteins mentioned in (*59,60*);
- Gamma secretase substrates: Proteins mentioned in (*61*).

### 2.6 Neuroimaging analysis

We studied neuroimaging data only from ADNI, because this study collected longitudinal scans. As a measure of brain atrophy we took cortical thickness data from 34 cortical areas from the longitudinal processing pipeline in Freesurfer version 4.3 for 1.5T T1-weighted MRI scans, and v5.1 for 3T T1-weighted MRI scans (http://adni.loni.usc.edu). As a measure of amyloid accumulation, we used the region-specific SUVR values for florbetapir binding assessed by PET imaging in 34 cortical areas as parcellated by Freesurfer v4.5.0 (*62*). As a measure of brain metabolism we analysed fluorodeoxyglucose (FDG)-PET scans (*63*), and we determined average glucose metabolism for brain areas according to the Desikaan-Killiany atlas (*64*) matching the cortical thickness areas for 3461 preprocessed scans that were available from ADNI (see http://adni.loni.usc.edu/methods/pet-analysis-method/pet-analysis/ for preprocessing details). Briefly, the Desikan-Killiany atlas was warped from standard space to subject-space using the subject-specific normalization parameters in SPM12. Normalization failed for n=24 scans, leaving a total of n=3437 scans. Voxelwise FDG signals greater than 3×10-6 were averaged within each brain area, and standardized to the average uptake in the vermis and brain stem.

### 2.7 BGenomic assessment

#### DNA extraction

For ADNI subjects, DNA was extracted from blood (n=1040) or cell lines (n=69). ADNI samples were genotyped using either the Illumina 2.5-M or the Illumina OmniQuad array and were retrieved online from http://adni.loni.cule.edu/ (*65*). For EMIF-AD individuals, DNA was extracted locally at the collection site (n=805) or from whole blood (n=148) using QIAamp DNA Blood Mini Kit (QIAGEN GmbH, Hilden, Germany) at the University of Lübeck. EMIF-AD MBD samples were genotyped using the Illumina Global Screening array (GSA) with shared custom content (Illumina, Incl) at the Institute of Clinical and Medical Biology (UKSH, Campus-Kiel). A total of 936 EMIF-AD MBD samples passed post-experiment QC.

#### Genotype imputation and quality control

Data processing and quality control was performed using GenomeStudio Software (v2.0.04, Illumina, Inc.), PLINK software (v1.9) and bcftools (v1.9), as described previously (*66*). Briefly, SNPs were excluded if they had a minor allele frequency (MAF) less than 1%, deviated significantly from Hardy-Weinberg equilibrium (HWE) (p<5×10-6) in the total sample of founder individuals, or had a call rate of less than 95%. We only used SNPs with no more than 2% genotype missingness and removed samples with excess heterozygosity rate (>5 SD). Individuals with sex mismatches were excluded from analysis. To identify ethnic outliers, a principal component analysis of ancestry (PCA) was performed based on 1000 Genomes clustering, phase 3 using PLINK) (*67*). Individuals of non-European descent were excluded from analysis. Relatedness was assessed through identity by descent, and family relations up to second degree were excluded. After filtering, the ADNI genotype data included 747 individuals and EMIF-AD 931 individuals. SNPs were locally imputed using Minimac 3 (*68*) to the Haplotype Reference Consortium (HRC) reference panel (*69*), leaving 4,583,773 SNPs (ADNI) and 7,778,465 SNPs (EMIF-AD) for final analyses. To account for population structure, principal components (PC1-PC20) were computed on a subset of relatively uncorrelated (r2<0.2) SNPs.

#### Classification of APOE genotypes

For ADNI subjects, APOE ε4 genotype was assessed with two SNPs; rs429358 the ‘ε4-allele’ and rs7412 the ‘ε2 allele’), using DNA extracted by Cogenics from a 3mL aliquot of EDTA blood. We used imputed DNA microarray genotype data available from ADNI-1, ADNI-GO, ADNI-2 processed with GenomeStudio v2009.1. APOE ε4 genotypes in EMIF-AD were generated as described elsewhere (*70*). Briefly, APOE ε4genotypes were determined either directly (rs7412) or by imputation (rs429358). For 80 samples for which no local *APOE* genotype was available, and for 45 mismatches between locally derived and GSA–derived genotypes (4.8%), APOE ε4 genotype was determined using TaqMan assays (ThermoFisher Scientific, Foster City, CA) on a QuantStudio-12K-Flex system in a 384-well format.

### 2.8 GWAS and post-GWAS analyses

Genome-wide association study (GWAS) analyses on CSF t-tau abnormality status in subjects with abnormal amyloid were performed separately in ADNI (a+t+=246, a+t-=238) and EMIF-AD (a+t+=294, a+t-=155) using PLINK software (V1.90). We used a logistic regression model including the PC1-PC3. Genome-wide significance was defined as p≤5e-08. Meta-analysis on ADNI and EMIF-AD GWAS summary statistics was performed using METAL (*71*). MAGMA software package was used to compute gene scores and geneset scores for biological pathways based on p-values of the meta-analysis summary statistics. We considered for further analysis gene scores based on at least 6 SNPs and geneset scores with information of at least 6 genes.

### 2.9 Polygenic and other genetic risk score analysis

Polygenic risk scores (PGRS) for AD were computed for each subject using PRSice (V2.3) (*72*). PGRS were calculated by adding the sum of each allele weighted by the strength of its association with AD risk as calculated previously by the largest GWAS on AD (71,880 cases; 383,378 controls) (*29*). AD cases were defined as patients clinically diagnosed with AD-type dementia or individuals with a parental history of AD (i.e., AD-by-proxy). AD-by-proxy showed strong genetic correlation with the clinical diagnosis of AD (rg = 0.81). Clumping was performed prior to calculating PGRS, to remove SNPs that are in LD (r2>0.1) within a slicing 1M bp window. After clumping, PGRS were computed using various SNP inclusion thresholds (i.e., p≤1e-30, p≤1e-8, p≤1e-5, p≤0.01, p≤0.02, p≤0.03, p≤0.04, p≤0.1, p≤0.2, p≤0.3, p≤0.4,p≤0.5). In order to explore how specific genetic alterations were associated with CSF proteomic profiles, we calculated for the top 3 genes and top 3 GO BP genesets from the MAGMA analysis, a selection of 9 GO-BPs that were associated with abnormal t-tau both in CSF enrichment analysis and MAGMA geneset analysis, and 2 GO-BP associated H3 acetylation, as described in the text, gene-specific risk scores (GRS) including only SNPs located on the respective genes with clumping as described above and computation of GRS using various SNP inclusion thresholds. We used weights from one cohort to generate GRS in the other cohort.

### 2.10 Statistics

Unless specified otherwise, we report on uncorrected p-values with statistical significance set at p<0.05.

#### Cross-sectional analysis

The group comparisons between AD subgroups and controls were performed using ANOVA correcting for age and gender.

#### Longitudinal analysis

Change in cognition and imaging markers were assessed using ADNI data with linear mixed models including as main terms group, time and the interaction group*time, and correcting for age and gender, and additionally for level of education for cognitive markers.

#### Protein change with disease severity

We investigated within the AD normal t-tau and AD abnormal t-tau groups whether proteins differed between stages by performing MANOVA tests. We calculated p for trend over disease severity categories. A protein was considered to decrease or increase with disease severity if the linear trend was statically significant or if there was a difference between controls and MCI, controls and dementia, or MCI and dementia and the trend analysis supported an increase or decrease with severity.

#### Genetic analysis

Analysis were performed in all individuals with GWAS data and information on abeta42 and t-tau status. All (P)GRS were compared between AD subgroups using linear models. Associations between GRS scores for risk genes and CSF protein levels were tested with linear models adjusted for age and sex. In order to reduce number of tests GRS scores were selected for SNP inclusion threshold that best differentiated within AD between abnormal and normal t-tau.

## Data Availability

Raw proteomic data has been deposited in the ProteomeXchange Consortium via the PRIDE partner repository with the dataset identifier 10.6019/PXD019910.

## Acknowledgements

This work has been supported by ZonMW Memorabel grant programme #73305056 (BMT) and #733050824 (BMT and PJV), the Swedish Research Council (#2018-02532, HZ), the European Research Council (#681712, HZ) and Swedish State Support for Clinical Research (#ALFGBG-720931, HZ), the Alzheimerfonden (Grant no. AF-930934) and Dtiftelsen Gamla tjänarinnor (JG), and the Innovative Medicines Initiative Joint Undertaking under EMIF grant agreement #115372 (PJV, HZ, SV, IB). Statistical analyses were performed at the VUmc Alzheimer Center that is part of the neurodegeneration research program of the Neuroscience Campus Amsterdam. EMIF-AD MBD proteomic analyses were performed at the Department of Psychiatry and Neurochemistry, the Sahlgrenska Academy at the University of Gothenburg, Sweden. The VUmc Alzheimer Center is supported by Stichting Alzheimer Nederland and Stichting VUmc fonds. HZ is a Wallenberg Academy Scholar. The Leuven cohort was funded by Stichting Alzheimer Onderzoek (#11020, #15005, #13007) and the Vlaamse Impulsfinanciering voor Netwerken voor Dementie-onderzoek (IWT #135043). The Lausanne cohort was supported by the Swiss National Research Foundation SNF (#320030_141179).

We thank Drs. Andre Franke and Michael Wittig as well as Mrs. Tanja Wesse and Sanaz Sedghpour Sabet for their help with the genome-wide genotyping experiments. We thank Dr. Fabian Kilpert for his help with the processing and management of the genome-wide genotyping data.

Data was used for this project of which collection and sharing was funded by the Alzheimer’s Disease Neuroimaging Initiative (ADNI) (National Institutes of Health Grant U01 AG024904) and DOD ADNI (Department of Defense award number W81XWH-12-2-0012). ADNI is funded by the National Institute on Aging, the National Institute of Biomedical Imaging and Bioengineering, and through generous contributions from the following: AbbVie, Alzheimer’s Association; Alzheimer’s Drug Discovery Foundation; Araclon Biotech; BioClinica, Inc.; Biogen; Bristol-Myers Squibb Company; CereSpir, Inc.; Cogstate; Eisai Inc.; Elan Pharmaceuticals, Inc.; Eli Lilly and Company; EuroImmun; F. Hoffmann-La Roche Ltd and its affiliated company Genentech, Inc.; Fujirebio; GE Healthcare; IXICO Ltd.; Janssen Alzheimer Immunotherapy Research & Development, LLC.; Johnson & Johnson Pharmaceutical Research & Development LLC.; Lumosity; Lundbeck; Merck & Co., Inc.; Meso Scale Diagnostics, LLC.; NeuroRx Research; Neurotrack Technologies; Novartis Pharmaceuticals Corporation; Pfizer Inc.; Piramal Imaging; Servier; Takeda Pharmaceutical Company; and Transition Therapeutics. The Canadian Institutes of Health Research is providing funds to support ADNI clinical sites in Canada. Private sector contributions are facilitated by the Foundation for the National Institutes of Health (www.fnih.org). The grantee organization is the Northern California Institute for Research and Education, and the study is coordinated by the Alzheimer’s Therapeutic Research Institute at the University of Southern California. ADNI data are disseminated by the Laboratory for Neuro Imaging at the University of Southern California.

## Author Contributions

Study design: BMT, PJV.

Design proteomics EMIF-AD: JG, HZ.

Design statistical analyses: BMT, PJV.

Data acquisition, processing, technical: JG, LR, IJ, ED, SH, VD, MT, FRJV, JP,

PMLA, RV, AL, JLM, SE, YFL, LF, KS, LB, SL, JS, SV, IB, DP, GS, KB, PS, CET, HZ, PJV.

Manuscript drafting: BMT, PJV.

Manuscript revising: all authors.

## Supplemental figures

**sFigure 1.**
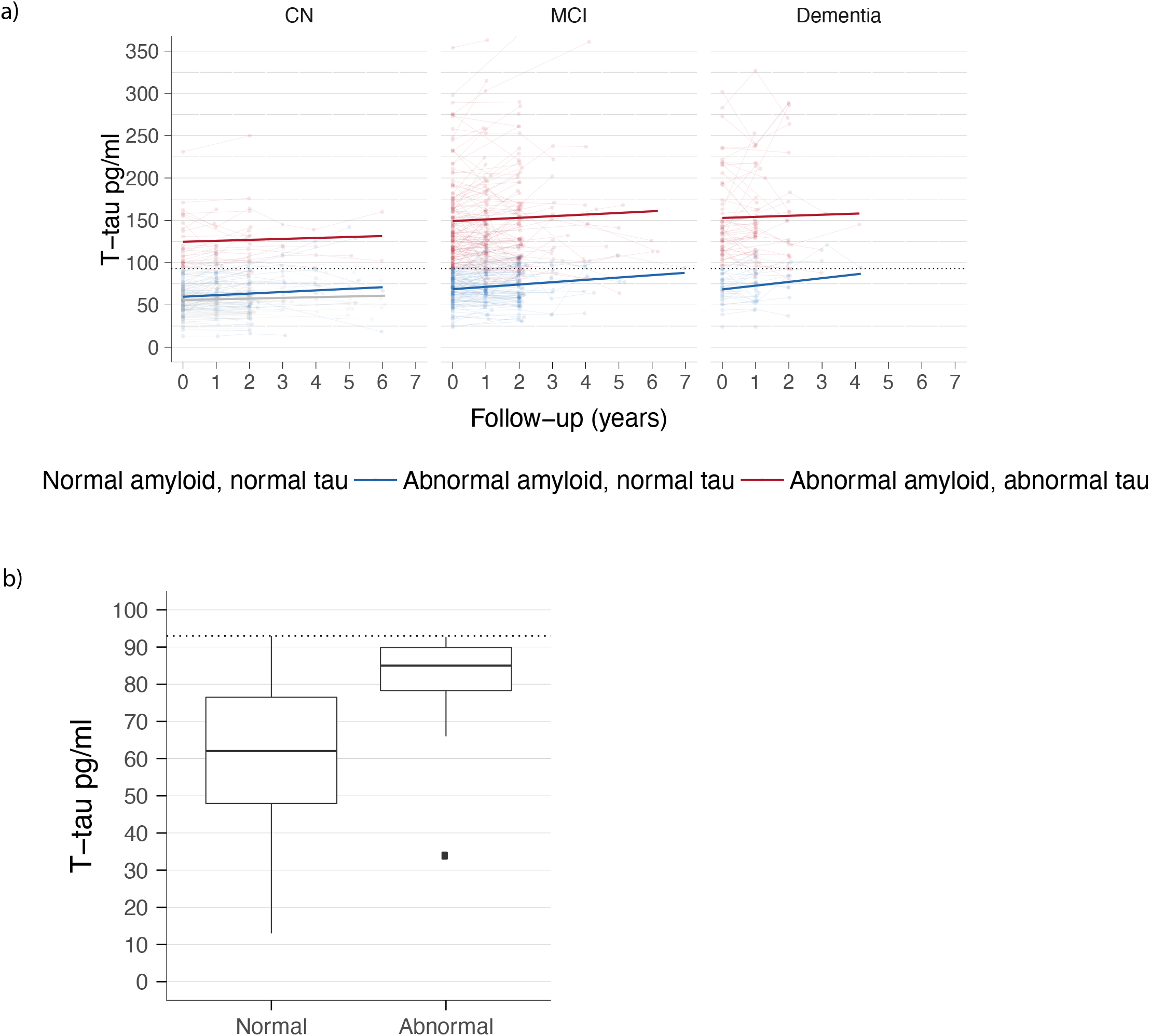
Longitudinal change in CSF total tau. A) Change in t-tau as function of group (control (grey), abnormal CSF Aβ_1-42_ and abnormal t-tau (red), and abnormal Aβ_1-42_ and normal t-tau (blue) and clinical stage. CN is normal cognition, MCI=mild cognitive impairment. B) Baseline t-tau concentration of individuals with normal t-tau at baseline according to t-tau status at last follow-up. Individuals with abnormal t-tau at follow-up scored just below the cut-point of abnormal t-tau (dotted line) at baseline.

**sFigure 2.**
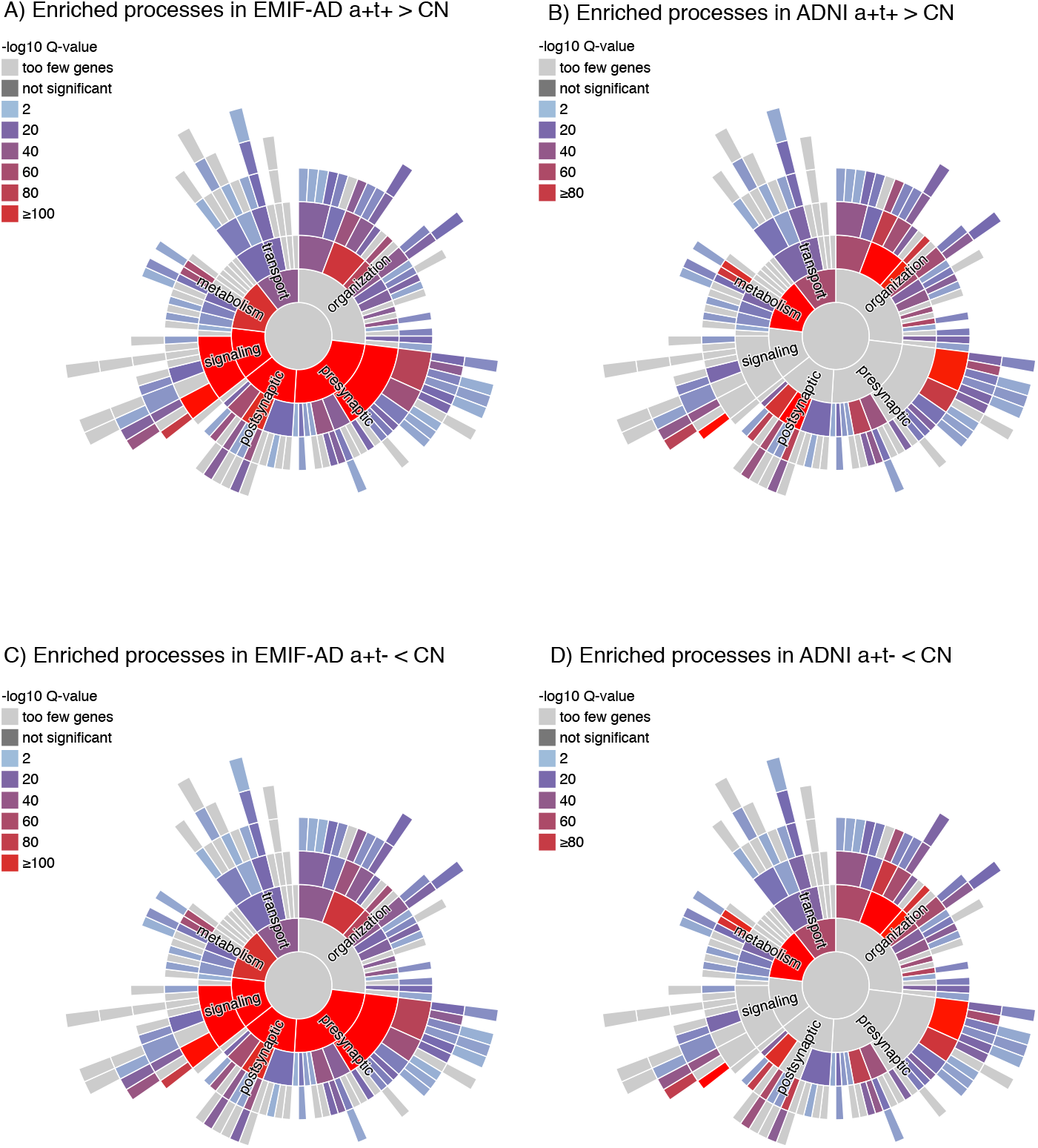
Enrichment of proteins for synaptic processes from SynGO. Enrichment of SynGO synaptic biological processes in EMIF-AD (A, C) and ADNI (B, D). Enriched processes in proteins with increased levels in individuals who have abnormal CSF Aβ_1-42_ and abnormal t-tau (a+t+) relative to control (CN, figure A, B) are similar to processes enriched in proteins with decreased level in individuals with abnormal CSF Aβ_1-42_ and normal t-tau (a+t-) relative to CN (figure C, D). Data are shown in sTable 3b.

**sFigure 3.**
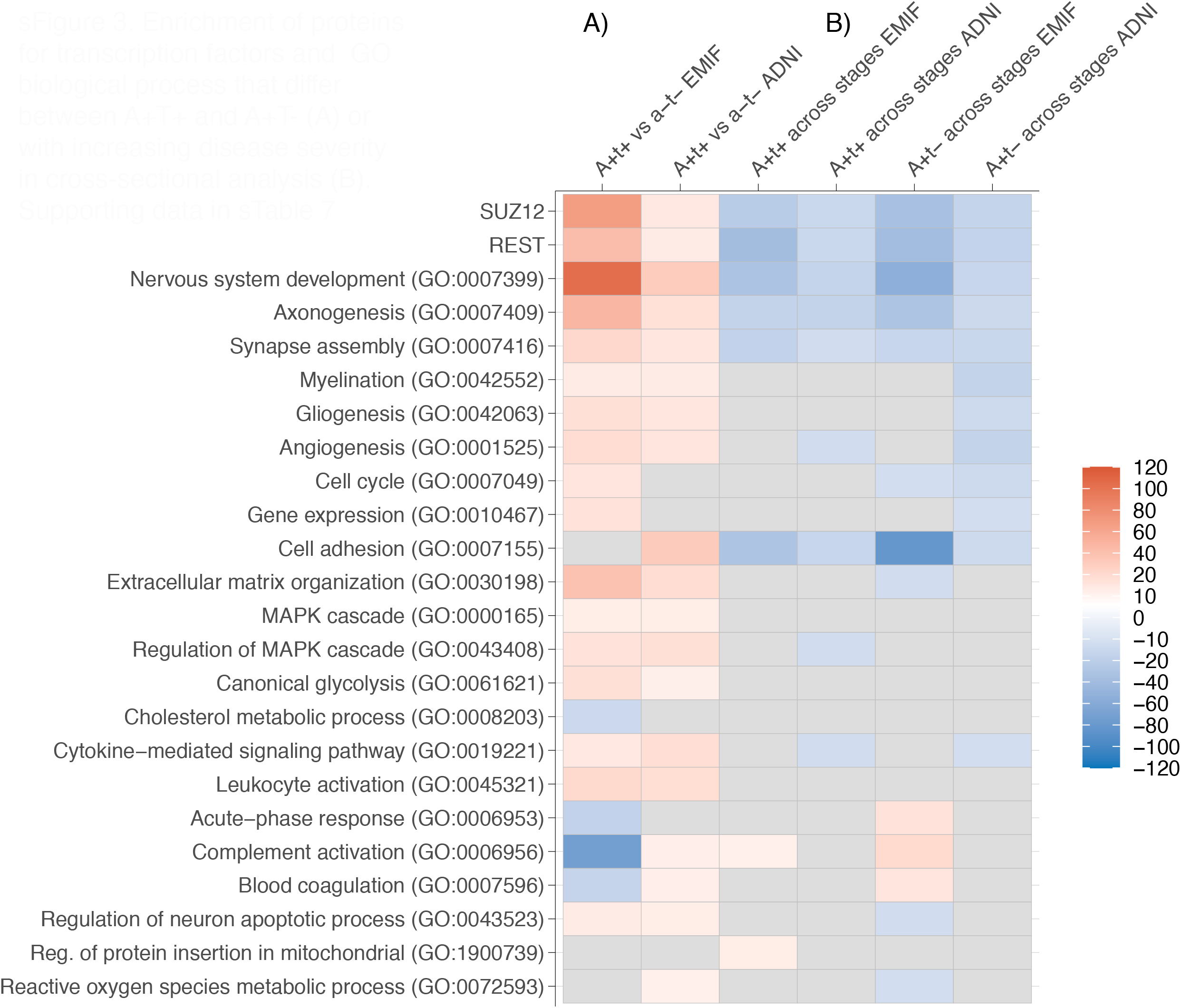
Enrichment of proteins for transcription factors and GO biological process. A) Heatmap showing p-value (log p-value*-1) of selected enriched GO biological processes and SUZ12 and REST transcription factors of proteins that were higher in in individuals with abnormal Aβ_1-42_ and t-tau (a+t+ (red)) or lower (blue) relative to individuals with abnormal Aβ_1-42_ and normal t-tau (a+t-). B) GO-BP associated with proteins that showed a decrease in concentration with disease stage (blue) or increase with disease stage (red). Data are shown for ADNI and EMIF-AD separately. Data used in heatmap are shown in sTable 7. P-values of all GO biological processes are listed in sTable 3a.

**sFigure 4.**
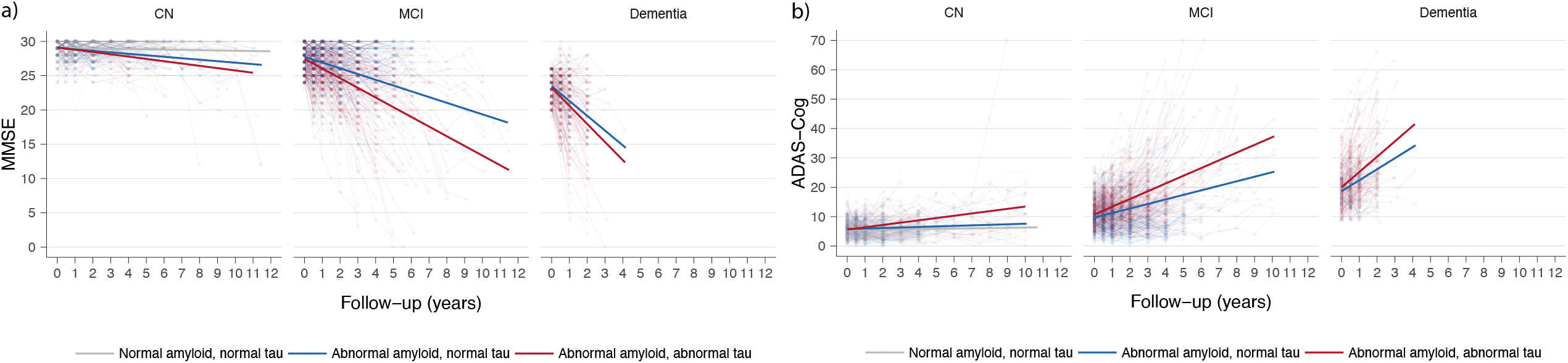
Longitudinal change in cognitive measures according to diagnostic group and clinical stage. MMSE (A) and ADAS-Cog (B). Change of controls are shown in grey, of individuals with abnormal Aβ_1-42_ and t-tau in red, and of individuals with abnormal Aβ_1-42_ and normal t-tau in blue. Data are shown in sTable 6a. Data are from ADNI only. CN=normal cognition; MCI=mild cognitive impairment; a+t+= abnormal Aβ_1-42_ and t-tau; a+t-= abnormal Aβ_1-42_ and normal t-tau.

**sFigure 5.**
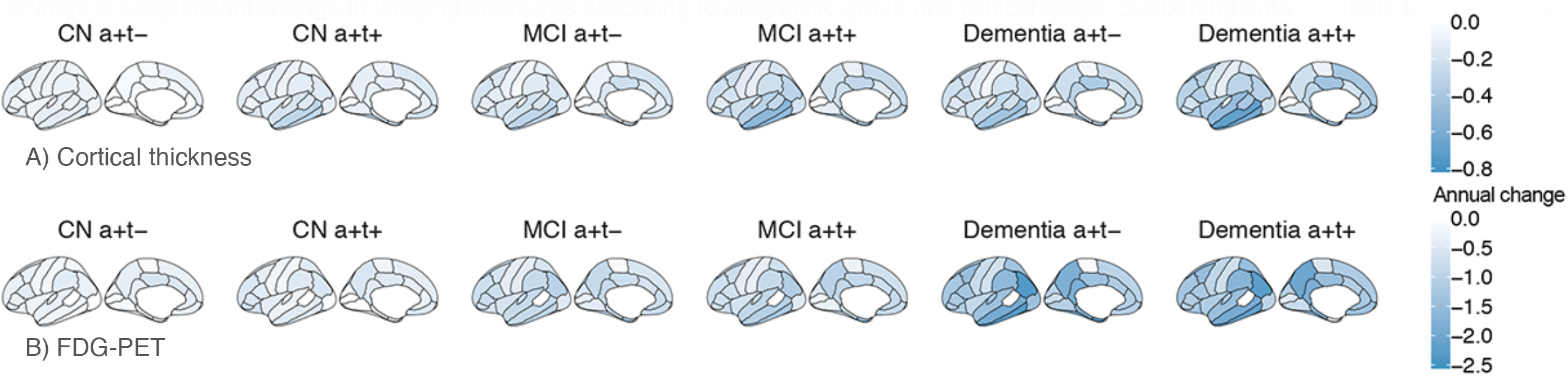
Longitudinal change in imaging measures according to diagnostic group and clinical stage. (A) cortical thickness and (B) FDG-PET glucose metabolism. Data are from ADNI only. Data are shown in sTable 6b. CN=normal cognition; MCI=mild cognitive impairment; a+t+= abnormal Aβ_1-42_ and t-tau; a+t-= abnormal Aβ_1-42_ and normal t-tau.

## Supplemental tables

**Supplemental table 1. Descriptives**

<u>Supplemental Table 1a.</u> Characteristics of individuals with information on CSF Aβ_1-42_ and t-tau at first visit.

<u>Supplemental Table 1b.</u> Characteristics of individuals with available CSF proteomic data.

<u>Supplemental Table 1c.</u> Aβ_1-42_ Cut-point definition.

**Supplemental table 2. Protein data**

<u>Supplemental Table 2a.</u> Number of proteins with change in concentration relative to reference or disease stage.

<u>Supplemental Table 2b.</u> Longitudinal t-tau data (ADNI only)

<u>Supplemental Table 2c.</u> Protein annotation and statistics of group comparisons.

<u>Supplemental Table 2d.</u> Cell-type specific proteins with largest differences between abnormal Aβ_1-42_ and t-tau and abnormal Aβ_1-42_ and normal t-tau.

<u>Supplemental Table 2e.</u> Proteins that differ in preclinical abnormal Aβ_1-42_ and t-tau or preclinical abnormal Aβ_1-42_ and normal t-tau from control group.

**Supplemental table 3. Protein enrichment analysis**

<u>Supplemental Table 3a.</u> Full list of GO biological processes associated with proteins that differ according to group or severity.

<u>Supplemental Table 3b.</u> SynGO enriched synaptic cellular components and biological processes that differ for proteins that are increased in abnormal Aβ_1-42_ and abnormal t-tau versus CN or decreased in abnormal Aβ_1-42_ and normal t-tau versus CN.

**Supplemental Table 4 Genetic analysis**

<u>Supplemental Table 4a.</u> Estimated marginal means (se) of AD GWAS-based polygenic risk scores in controls, abnormal Aβ_1-42_ and normal t-tau and abnormal Aβ_1-42_ and abnormal t-tau in total sample and according to clinical stage.

<u>Supplemental Table 4b.</u> Top 1000 SNPS from GWAS on abnormal Aβ_1-42_ and normal t-tau versus abnormal Aβ_1-42_ and abnormal t-tau in pooled ADNI and EMIF-AD cohorts.

<u>Supplemental Table 4c.</u> Difference in MAGMA gene score between abnormal Aβ_1-42_ and abnormal t-tau and abnormal Aβ_1-42_ and normal t-tau based on GWAS in pooled ADNI and EMIF-AD cohorts.

<u>Supplemental Table 4d.</u> Difference in GO biological process MAGMA geneset score between abnormal Aβ_1-42_ and abnormal t-tau and abnormal Aβ_1-42_ and normal t-tau based on GWAS in pooled ADNI and EMIF-AD cohorts.

**Supplement Table 5. Correlation between genetic risk score and CSF protein concentrations**

<u>sTable 5a.</u> Summary data on correlation of selected genetic risk scores with CSF protein correlations

<u>Supplemental Table 5b.</u> Correlation between genetic risk score and CSF protein level in individuals with abnormal Aβ_1-42_.

**Supplemental Table 6. Longitudinal analysis of cognitive and imaging outcomes**

<u>Supplemental Table 6a.</u> Cross-sectional and annual change effects on clinical measures

<u>Supplemental Table 6b.</u> Cross-sectional and annual change effects of imaging measures. Data from ADNI only.

**Supplemental Table 7. Data used in heatmap in figure 2 and sFigure 3**.

